# Bridging Adults to Heart Transplant: Temporary versus Durable Mechanical Circulatory Support and Post-Transplant Outcomes

**DOI:** 10.64898/2025.12.08.25341866

**Authors:** Abhishek Jaiswal, William L. Baker, Suguru Ohira, Dina Al-Rameni, Sooyun Caroline Tavolacci, David A. Baran

## Abstract

**Background:** The current United States donor heart allocation system prioritizes patients receiving temporary mechanical circulatory support over those with durable left ventricular assist devices (d-LVADs), but the impact on post-transplant survival remains unclear.

**Objectives:** To evaluate post-transplant outcomes in patients bridged with d-LVAD versus t-LVAD before and after the 2018 United Network for Organ Sharing (UNOS) allocation policy change.

**Methods:** Using the UNOS database, we analyzed 24,795 adult first-time HT recipients from 2011–2023, stratified by device type at transplant: d-LVAD (43.3%), t-LVAD (6.4%), or no LVAD. Outcomes included survival at 30 days, 90 days, 1 year, and 2 years. Risk-adjusted analyses were performed using Cox proportional hazards models. Subgroup analysis examined time on LVAD and the impact of organ preservation on outcomes.

**Results:** Compared to t-LVAD and no-LVAD recipients, d-LVAD recipients had significantly higher adjusted mortality rates at all time points (hazard ratios ranged from 1.44 at 30 days to 1.18 at 2 years; p < 0.001). The mortality gap was more pronounced under the current allocation era. In patients with device duration data, ≥2 years on LVAD was associated with a 39% higher 1-year mortality risk (HR 1.39, 95% CI 1.15–1.68). No significant differences in 1-year mortality were observed between DCD donor and machine-perfused donor transplant subgroups by LVAD status.

**Conclusions:** Post-transplant survival is worse with d-LVAD bridging, particularly under current allocation rules, and prolonged LVAD support further elevates risk. These findings underscore the need to reevaluate LVAD strategy and transplant prioritization, considering evolving allocation policies.

**Condensed Abstract:** In a UNOS analysis of 24,795 heart transplants (2011–2023), patients bridged with durable LVADs (43.3%) consistently experienced worse post-transplant survival than those with temporary LVADs (6.4%) or no device. Adjusted mortality was higher at all time points (HR 1.44 at 30 days to 1.18 at 2 years; p<0.001), with disparities becoming more pronounced after the 2018 allocation change. Prolonged LVAD use (≥2 years) further increased the risk. These findings highlight the need to reevaluate durable LVAD strategies and transplant prioritization.

**Preprint Server:** None

## INTRODUCTION

Advances in technology, including smaller devices, longer battery life, and a deeper understanding of the pathophysiology of circulation in patients with durable left ventricular assist devices (d-LVADs), have contributed to a relatively low mortality risk for clinically stable patients. Recognizing this, contemporary donor heart allocation in the United States prioritizes heart transplantation (HT) for sicker populations supported with temporary LVAD devices (t-LVAD) over stable patients waiting at home with a stable d-LVAD support and no significant device complications. Due to this change in organ allocation, the number of d-LVADs implanted as a bridge to transplant (BTT) in the current allocation era has decreased significantly when compared to the prior allocation era.[1] However, patients listed as the United Network for Organ Sharing (UNOS) status 2 with t-LVAD have been allowed to wait only two weeks. After this period, an extension submission is required to justify the ongoing wait as a priority status due to ineligibility for LVAD, which must be submitted on an ongoing basis if they don’t receive HT. This decision becomes complicated when patients are optimal candidates for both HT and LVAD implantation, yet transplant selection committees must determine the most appropriate therapy. Often, there is a default preference for LVAD implantation to avoid overburdening the HT waitlist and to adhere to guidelines established by the organ allocation scheme.

Though this approach optimizes organ allocation, it may limit patient autonomy and poses challenges for providers due to unclear and conflicting data on post-transplant survival in d-LVAD versus non-LVAD patients.[2–9] Additionally, optimal timing between d-LVAD implantation and HT remains controversial, with some studies reporting poor outcomes regardless of time spent on d-LVAD support.[2,4,6,8] Long waiting times for HT often lead to the occurrence of serious device-related complications before an organ becomes available.[10] In the current donor allocation system in the United States, 74% of BTT d-LVAD recipients did not receive HT until complications qualifying for a higher status occurred.[11] This observation is particularly relevant due to the possibility of inferior post-transplant outcomes in patients with d-LVAD-associated complications.[12,13] Moreover, despite all the necessary measures and evaluation of the right heart before d-LVAD implantation, right ventricular failure (RVF) remains a serious complication after LVAD implantation. Notably, RVF contributes significantly to postprocedural morbidity and mortality as well as eligibility for transplantation due to RVF failure-related end-organ damage.[14] Hence, concerns around the impact of adverse events, end-organ dysfunction, and increased procedural complexity-such as prior sternotomy and longer cardiopulmonary bypass time during transplant surgery-on survival after HT remain. Additionally, these complications may potentially interfere with HT eligibility. In addition, d-LVAD patients may experience psychological distress after implantation, which can affect the patient’s overall health, BTT strategy, and clinical outcome.[15] However, a recent comparative analysis of data from the MOMENTUM 3 trial and UNOS registry reported similar two-year survival rates among younger patients who received either LVAD or HT, concluding d-LVAD as a BTT could offer enhanced net prolongation of life, especially in the younger patients who generally have fewer comorbidities, longer life expectancy, and greater physiological resilience (16). By contrast, t-LVAD as a BTT provides comparable or even superior outcomes post-HT despite being used in a sicker patient population, challenging the default preference for d-LVAD as the BTT strategy. [17,18] Therefore, it is pivotal to better understand the clinical outcomes of patients who receive an LVAD as a bridge to HT in the contemporary practice.

In this evolving clinical and policy backdrop, we analyzed a large multicenter national database from the United States, examining a contemporary patient population before and after changes to the allocation system. Our study aimed to evaluate the short- and mid-term clinical outcomes of patients who underwent BTT with d-LVAD versus t-LVAD at the time of HT. We hypothesized that bridging with t-LVAD would provide superior outcomes, and that outcomes following BTT with d-LVAD would be inferior under the current allocation system compared to the prior era due to reduced access to donor organs.

## METHODS

### Data Source and Study Population

The United Network for Organ Sharing database, collected by the Organ Procurement and Transplantation Network, was explored. All adults (≥18 years old) who underwent their first heart-only transplantation from January 1st, 2011, to December 31^st^, 2023, were included. Follow-up data was available through December 31^st^, 2024. Individuals listed for dual-organ transplantation or re-transplantation, as well as those who were on either extracorporeal membrane oxygenation, intra-aortic balloon pump, or a bi-ventricular assist devices were excluded. The database contains de-identified patient-level records, including recipients, donors, and candidates on the wait list. As this study utilized a de-identified dataset from the UNOS registry, which does not include patient or center identifiers, it was exempt from Institutional Review Board approval.

Patients were categorized into three groups: Those who had a d-LVAD at the time of transplant (HeartMate 2^TM^, Abbott, Abbott Park, IL; HeartMate 3^TM^, Abbott, Abbott Park, IL; or HVAD^TM^, Medtronic, Minneapolis, MN); those who had a t-LVAD at the time of transplant (Impella^TM^ devices, Abiomed, Danvers, MA-Impella 5.0^®^, Impella 5.5^®^, Impella 2.5^®^, Impella CP^®^, or Centrimag^TM^, Abbott Medical, Abbott Park, IL ); those who had no LVAD at the time of transplant.

### Patient Outcomes

Baseline characteristics were collected at the time of organ placement for donors and at the time of listing or transplant for recipients. The outcomes of interest included 30-day, 90-day, 1-year, and 2-year survival after HT. We also examined the cause of death, including multiorgan failure, infectious, cardiovascular, pulmonary, cerebrovascular, hemorrhagic, or other causes. We also evaluated the impact of the organ allocation era by comparing outcomes before and after the 2018 UNOS heart allocation policy change. We analyzed the effect of time on LVAD support, both as less than or greater than 2 years, and in four quartiles, on 1-year post-transplant mortality. Additionally, given the increasing use of donation after circulatory death (DCD) and donation after brain death (DBD) with machine perfusion (MP) HT since 2019, we examined outcomes of transplants in our study cohort involving DCD and DBD using MP retrieval.

### Statistical Analyses

All demographics and clinical variables of HT recipients stratified by LVAD use are summarized with percentages for categorical variables and median with interquartile ranges (25^th^ and 75th percentile) for continuous variables. We compared baseline characteristics between groups using either Pearson’s chi-square for categorical or the Wilcoxon rank sum test for continuous variables. Kaplan-Meier survival analysis with log-rank test (global and pairwise) compared the time from organ listing to death between groups at each desired time point.

We used multiple imputations with chained equations, assuming missingness to be random, using predictive mean matching with the “mice” package (version 3.16.0) in R. [19] All predictors and the outcomes were included in the imputation model. Because the missingness rate of previous cardiac surgery was approximately 2%, we generated three multiply imputed datasets, with convergence examined through visual inspection of plots and diagnostics. Analyses were performed on each imputed dataset and pooled using Rubin’s rules.

To evaluate risk-adjusted mortality between the UNOS eras in those who received induction therapy, a Cox proportional hazards model was created using the ‘rms’ package (version 6.9-0) in R. Characteristics shown to be associated with post-transplant mortality were used in the models and included recipient age, body mass index (BMI), male gender, serum creatinine, serum bilirubin, cold ischemic time, ischemic heart failure etiology, history of diabetes mellitus, prior cardiac surgery, donor age, donor-recipient predicted heart mass ratio [20], donor-recipient gender mismatch, and center effects with average number of transplants per year. Continuous variables were modeled with restricted cubic splines (3 knots). Results are reported as adjusted hazard ratio (HR) with corresponding 95% confidence interval (CI).

### Subgroup Analyses

To evaluate the robustness of our findings, several subgroup analyses were conducted. The first was conducted, whereby mortality at each time point was assessed separately before and after October 18^th^, 2018, the date of the UNOS heart allocation policy change. To further assess the impact of time on LVAD (with any included device) on mortality outcomes, we limited the cohort to only those who had either a d-LVAD or surgically implanted t-LVAD at the time of transplant and an available date of device implant. For those without an available date of device explant, we assumed it to be the date of transplant. Cox proportional hazards models for 1-year post-transplant mortality were constructed with time on LVAD modeled using a restricted cubic spline with 4 knots. We then categorized time on LVAD (for any included device) in two ways. The first was looking at individuals on LVAD for < or ≥ 2 years. For the second, we categorized individuals into time on LVAD quartiles as follows: 0-184 days, 185-493 days, 494-992 days, and 993+ days.

Additional subgroup analyses were performed, limiting the durable LVAD group to only those who had received a HM3 device. To evaluate the impact of listing status on 1-year mortality, analyses were stratified by final listing status with recipients designated as either high-priority (old policy status 1A and new policy status 1-3) or low-priority (all other status). We also compared the adjusted 1-year mortality risk between the main study groups for transplants with DCD and DBD transplants that utilized MP. Transplants with DCD were identified using the ‘NON_HRT_DON’ variable, while those utilizing MP were identified using the ‘HEART_PERFUSION’ variable, according to the UNOS data dictionary.

We performed data management using SAS 9.4 (SAS Institute, Cary, NC) and data analysis using R 4.4.3 (The R Foundation, https://www.r-project.org/) in RStudio (version 2025.05.0), with a p-value < 0.05 considered statistically significant. This report was written following the Strengthening in Reporting of Observational Studies in Epidemiology (STROBE) statement.[21]

## RESULTS

### Study Cohort

Between 2011 and 2023, a total of 24,795 individuals met study inclusion criteria (**Figure 1**). Of the 10,729 (43.3%) in the d-LVAD group, 5,291 (49.3%) had received a HeartMate 2^TM^, 2,222 (20.7%) received a HeartMate 3^TM^, and 3,216 (30.0%) received a HeartWare^TM^ HVAD^TM^. Of the t-LVAD group, most (92.7%) received an Impella^®^ device, with 379 (42.3%) being Impella 5.0 and 372 (41.5%) being the Impella 5.5, while the rest (n=65, 7.3%) received a CentriMag^TM^.

**Figure 1:**
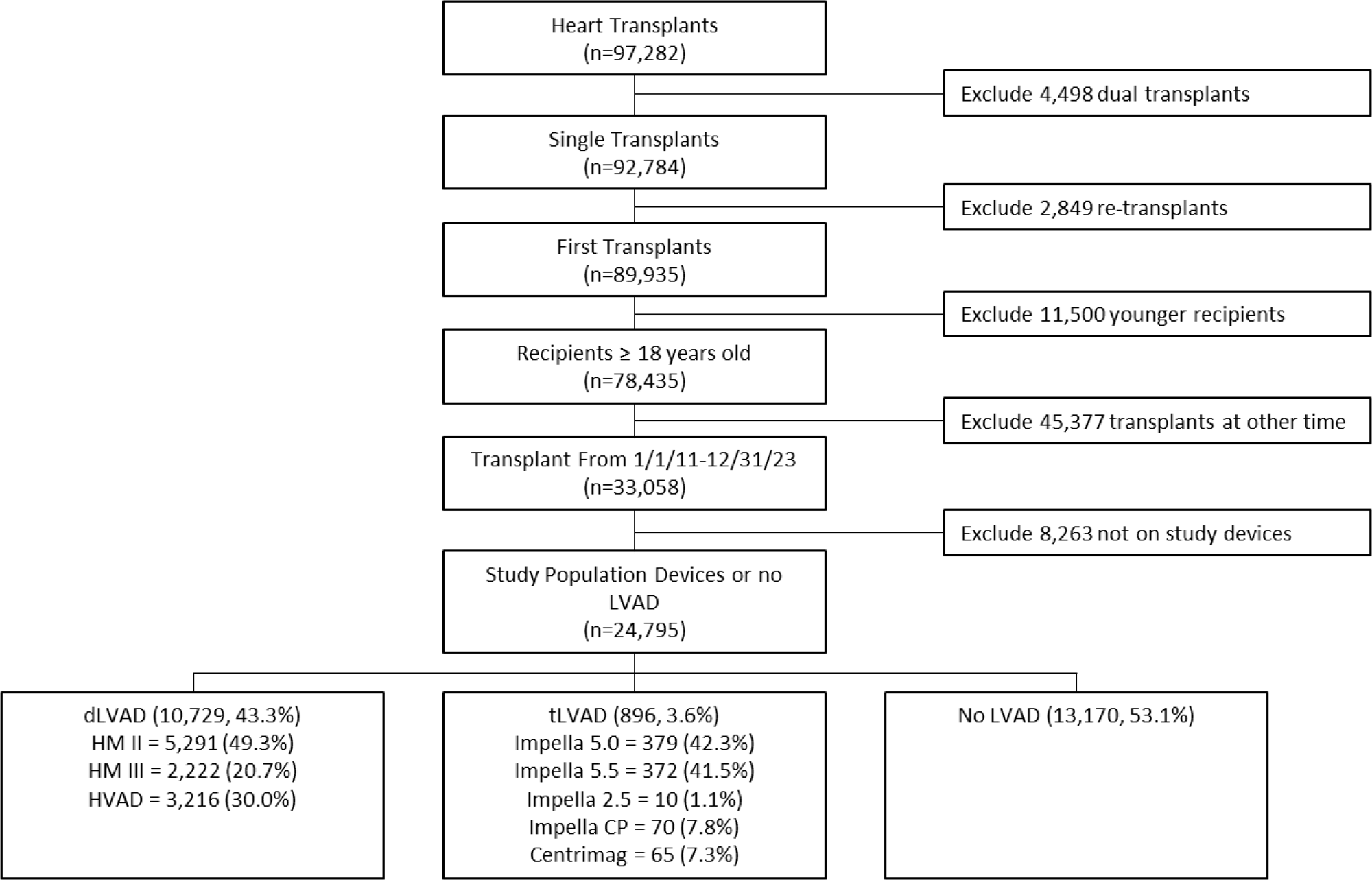
Study Cohort Selection Flow Diagram. d-LVAD = durable left ventricular assist device; HM = HeartMate; HVAD = Heartware Ventricular Assist Device; t-LVAD = temporary LVAD.

**Table 1** summarizes the baseline characteristics of the study cohort. Overall, the median (25^th^, 75^th^ percentile) age was 57 (47, 64) years, 72% were male, 64% were White, the median BMI was 27.6 kg/m^2^ (24.2, 31.4), and 51% had dilated cardiomyopathy. Compared to those with an t-LVAD, those with d-LVAD were significantly more likely to have a higher median BMI (29.0 kg/m^2^ [25.6, 32.6] vs 26.6 [23.7, 30.4]; p<0.001), less often had prior cardiac surgical procedures (44% vs. 78%; p<0.001), more often were listed as status 1A under the prior UNOS allocation system (45% vs. 10%) and status 3 (39% vs. 0.4%) under the new allocation system. Compared to those with a d-LVAD, those with a t-LVAD were younger (median age 56 vs. 57 years; p<0.001), were more often on a ventilator (3.2% vs. 0.3%; p<0.001), had higher pulmonary capillary wedge pressures (22 vs. 14 mmHg; p<0.001) and lower cardiac index (2.14 vs. 2.30 L/min/m^2^; p<0.001), more often listed as status 1 (10% vs. 4.8%) or 2 (89% vs. 23%) in the new UNOS system, and had a shorter median follow-up time (1.2 vs. 5.2 years; p<0.001). Center volume, measured as average number of transplants per year, differed between groups with the t-LVAD group being the lowest (18 [12, 27]) compared with the d-LVAD (21 [14, 27]) and no-LVAD groups (23 [15, 35]; p<0.001)

**Table 1.** Characteristics of the Study Cohort.

| **Characteristic** | **Total Cohort**<br>**(n=24,795)** | **No LVAD**<br>**(n=13,170)** | **Durable**<br>**LVAD**<br>**(n=10,729)** | **Temporary**<br>**LVAD (n=896)** | **p-value** |
| --- | --- | --- | --- | --- | --- |
| **Recipient Characteristics** |  |  |  |  |  |
| Age, years | 57 (47, 64) | 57 (46, 64) | 57 (47, 63) | 56 (45, 64) | 0.092 |
| Men | 17,905 (72%) | 8,663 (66%) | 8,518 (79%) | 724 (81%) | <0.001 |
| Race |  |  |  |  | <0.001 |
| Asian | 872 (3.5%) | 536 (4.1%) | 289 (2.7%) | 47 (5.2%) |  |
| Black | 5,399 (22%) | 2,436 (18%) | 2,748 (26%) | 215 (24%) |  |
| Other | 2,569 (10%) | 1,441 (11%) | 996 (9.3%) | 132 (15%) |  |
| White | 15,955 (64%) | 8,757 (66%) | 6,696 (62%) | 502 (56%) |  |
| BMI, kg/m<sup>2</sup> | 27.6 (24.2,<br>31.4) | 26.6 (23.2,<br>30.3) | 29.0 (25.6,<br>32.6) | 26.6 (23.7,<br>30.4) | <0.001 |
| HF Etiology |  |  |  |  | <0.001 |
| CHD | 927 (3.7%) | 846 (6.4%) | 77 (0.7%) | 4 (0.4%) |  |
| DCM | 12,651 (51%) | 5,811 (44%) | 6,289 (59%) | 551 (61%) |  |
| HCM | 790 (3.2%) | 708 (5.4%) | 77 (0.7%) | 5 (0.6%) |  |
| ICM | 7,286 (29%) | 3,393 (26%) | 3,633 (34%) | 260 (29%) |  |
| Other | 2,155 (8.7%) | 1,551 (12%) | 549 (5.1%) | 55 (6.1%) |  |
| RCM | 986 (4.0%) | 861 (6.5%) | 104 (1.0%) | 21 (2.3%) |  |
| Blood Type |  |  |  |  | <0.001 |
| A | 9,976 (40%) | 5,600 (43%) | 4,051 (38%) | 325 (36%) |  |
| AB | 1,398 (5.6%) | 963 (7.3%) | 403 (3.8%) | 32 (3.6%) |  |
| B | 3,767 (15%) | 2,124 (16%) | 1,521 (14%) | 122 (14%) |  |
| O | 9,654 (39%) | 4,483 (34%) | 4,754 (44%) | 417 (47%) |  |
| History of diabetes mellitus | 7,069 (29%) | 3,424 (26%) | 3,370 (31%) | 275 (31%) | <0.001 |
| Prior cardiac surgery | 19,988 (81%) | 8,585 (67%) | 10,729 (100%) | 674 (78%) | <0.001 |
| Ventilator at transplant | 126 (0.5%) | 69 (0.5%) | 28 (0.3%) | 29 (3.2%) | <0.001 |
| Creatinine, mg/dL | 1.17 (0.93, 1.41) | 1.14 (0.90, 1.40) | 1.20 (0.97, 1.43) | 1.10 (0.89, 1.41) | <0.001 |
| Bilirubin, mg/dL | 0.70 (0.40, 1.00) | 0.70 (0.46, 1.10) | 0.60 (0.40, 0.90) | 0.80 (0.51, 1.30) | <0.001 |
| PRA, % | 0 (0, 11) | 0 (0, 8) | 0 (0, 17) | 0 (0, 22) | <0.001 |
| PCWP, mmHg | 16 (10, 22) | 17 (11, 24) | 14 (9, 20) | 22 (16, 28) | <0.001 |
| CI, L/min/m<sup>2</sup> | 2.21 (1.85, 2.64) | 2.14 (1.78, 2.58) | 2.30 (1.95, 2.71) | 2.07 (1.66, 2.65) | <0.001 |
| Listing Status |  |  |  |  | <0.001 |
| 1A | 8,982 (36%) | 4,011 (30%) | 4,881 (45%) | 90 (10%) |  |
| 1B | 5,384 (22%) | 3,281 (25%) | 2,099 (20%) | 4 (0.4%) |  |
| 2 | 625 (2.5%) | 623 (4.7%) | 2 (<0.1%) | 0 (0%) |  |
| New Listing Status |  |  |  |  | <0.001 |
| 1 | 372 (3.8%) | 110 (2.1%) | 179 (4.8%) | 83 (10%) |  |
| 2 | 3,310 (34%) | 1,722 (33%) | 873 (23%) | 715 (89%) |  |
| 3 | 2,453 (25%) | 973 (19%) | 1,477 (39%) | 3 (0.4%) |  |
| 4 | 2,865 (29%) | 1,648 (31%) | 1,216 (32%) | 1 (0.1%) |  |
| 5 | 11 (0.1%) | 11 (0.2%) | 0 (0%) | 0 (0%) |  |
| 6 | 793 (8.1%) | 791 (15%) | 2 (<0.1%) | 0 (0%) |  |
| **Donor Characteristics** |  |  |  |  |  |
| Donor age, years | 31 (24, 40) | 32 (24, 41) | 31 (23, 40) | 31 (24, 39) | 0.015 |
| Donor Male | 17,121 (69%) | 8,294 (63%) | 8,159 (76%) | 668 (75%) | <0.001 |
| Donor BMI, kg/m<sup>2</sup> | 26.7 (23.4, 30.9) | 26.2 (23.0, 30.5) | 27.1 (23.9, 31.2) | 27.4 (23.9, 31.6) | <0.001 |
| Donor history of diabetes mellitus | 994 (4.0%) | 535 (4.1%) | 421 (4.0%) | 38 (4.3%) | 0.8 |
| Donor history of hypertension | 3,998 (16%) | 2,206 (17%) | 1,658 (16%) | 134 (15%) | 0.015 |
| **Transplant Characteristics** |  |  |  |  |  |
| Average center volume per year | 22 (14, 32) | 23 (15, 35) | 21 (14, 27) | 18 (12, 27) | <0.001 |
| Ischemic time, h | 3.28 (2.52, 3.92) | 3.27 (2.52, 3.92) | 3.28 (2.50, 3.93) | 3.60 (2.98, 4.18) | <0.001 |
| Donor-recipient PHM ratio | 1.12 (1.01, 1.24) | 1.12 (1.01, 1.25) | 1.11 (1.01, 1.23) | 1.12 (1.02, 1.23) | <0.001 |
| Recipient-Donor Mismatch | 5,582 (23%) | 3,355 (25%) | 2,041 (19%) | 186 (21%) | <0.001 |
| Follow-up Time, y | 5.0 (2.0, 8.0) | 5.0 (2.0, 8.1) | 5.2 (2.2, 8.0) | 1.2 (1.0, 3.1) | <0.001 |
| Geographic Region |  |  |  |  | <0.001 |
| Midwest | 5,565 (22%) | 2,535 (19%) | 2,926 (27%) | 104 (12%) |  |
| Northeast | 5,576 (22%) | 2,724 (21%) | 2,681 (25%) | 171 (19%) |  |
| South | 8,659 (35%) | 4,786 (36%) | 3,424 (32%) | 449 (50%) |  |
| West | 4,995 (20%) | 3,125 (24%) | 1,698 (16%) | 172 (19%) |  |

Data are presented either as n (%) or median (25<sup>th</sup>, 75<sup>th</sup> percentile) and compared using either Pearson's chi-squared test or Wilcoxon rank sum test, respectively. BMI = body mass index; CHD = congenital heart disease; CI = cardiac index; DCM = dilated cardiomyopathy; HCM = hypertrophic cardiomyopathy; ICM = ischemic cardiomyopathy; LVAD= left ventricular assist device; PCWP = pulmonary capillary wedge pressure; PHM = predicted heart mass; PRA = panel-reactive antibody; RCM = restrictive cardiomyopathy.

### Post-Transplant Mortality

The unadjusted Kaplan-Meier survival curves for each time point are shown in **Figure 2**. The overall post-transplant survival at each timepoint (**Table 2**) was: 96.7% in 30 days; 94.6% in 90 days; 91.4% in 1 year; and 88.4% in 2 years. Under the old UNOS allocation era (before October 2018), the survival rates were as follows: 96.6% at 30 days, 94.5% at 90 days, 91.4% at 1 year, and 88.3% at 2 years. Under the current UNOS allocation era (after October 2018), the survival rates were 96.9% at 30 days, 94.7% at 90 days, 91.3% at 1 year, and 88.6% at 2 years.

**Figure 2:**
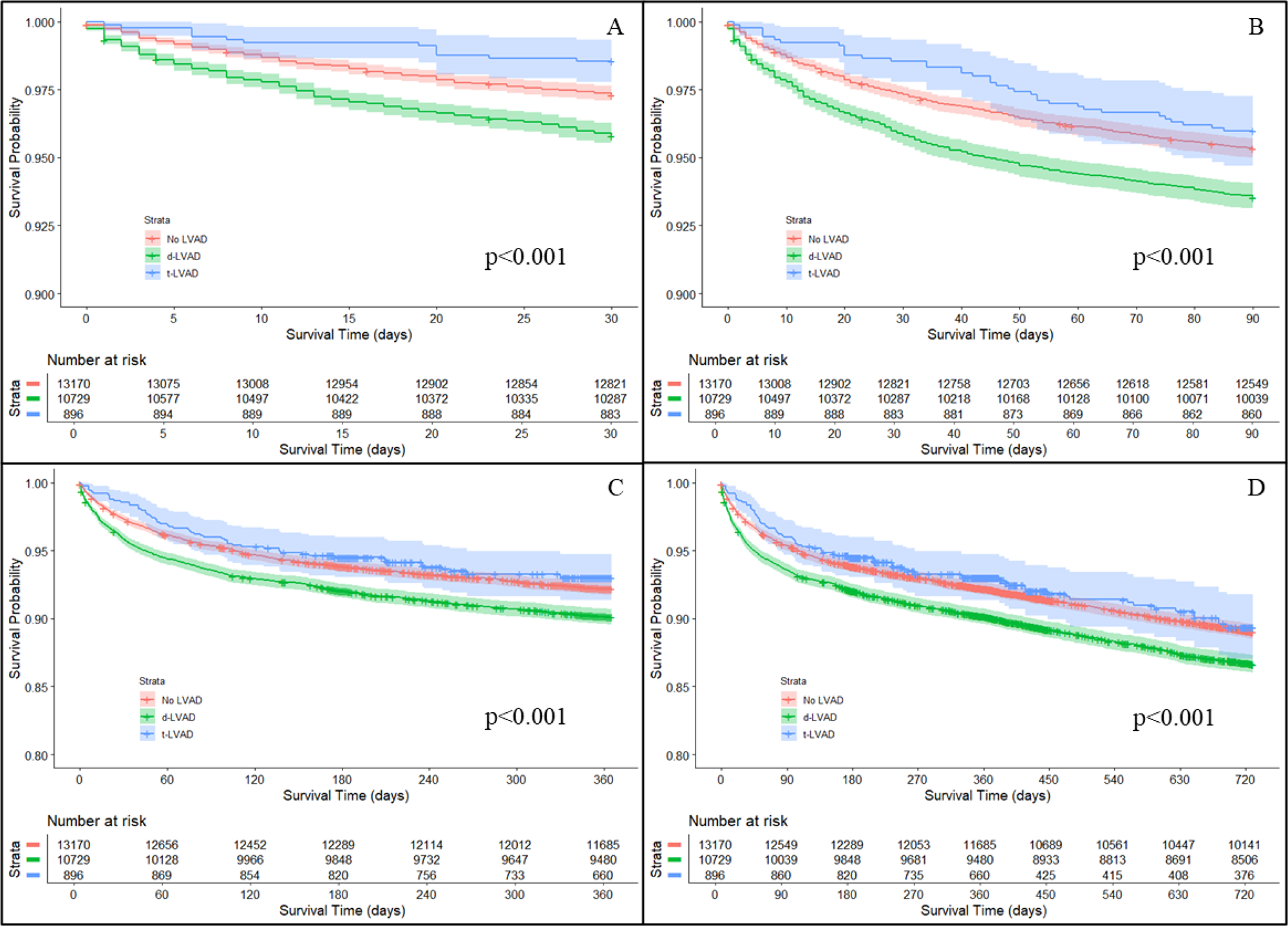
Kaplan-Meier Analysis for 30-Day (A), 90-Day (B), 1-Year (C), and 2-Year (D). LVAD = left ventricular assist device; d-LVAD = durable-LVAD; t-LVAD = temporary LVAD

**Table 2.** Survival Analysis of Study Groups.

| **Outcome** | **Survival Proportion (95% CI)** |  |  | **Multivariable Cox Proportional Hazard Regression Model** |  |  |
| --- | --- | --- | --- | --- | --- | --- |
|  | **No LVAD**<br>**(n=13,170)** | **Durable LVAD**<br>**(n=10,729)** | **Temporary LVAD**<br>**(n=896)** | **Durable LVAD vs.**<br>**no LVAD** | **Temporary LVAD vs.**<br>**no LVAD** | **Durable LVAD vs.**<br>**Temporary LVAD** |
| **30-Day** | 97.3% (97.0%, 97.6%) | 95.8% (95.4%, 96.2%) | 98.5% (97.5%, 99.2%) | HR 1.44 (1.24, 1.68) | HR 0.45 (0.26, 0.79) | HR 3.21 (1.82, 5.65) |
| **90-Day** | 95.3% (95.0%, 95.7%) | 93.6% (93.1%, 94.0%) | 96.0% (94.5%, 97.2%) | HR 1.28 (1.14, 1.44) | HR 0.71 (0.50, 0.99) | HR 1.83 (1.29, 2.59) |
| **1-Year** | 92.2% (91.8%, 92.7%) | 90.1% (89.5%, 90.7%) | 93.2% (91.3%, 94.8%) | HR 1.21 (1.10, 1.33) | HR 0.74 (0.57, 0.97) | HR 1.64 (1.25, 2.14) |
| **2-Year** | 89.4% (88.9%, 89.9%) | 86.9% (86.2%, 87.5%) | 91.3% (89.3%, 93.1%) | HR 1.18 (1.09, 1.28) | HR 0.80 (0.63, 1.01) | HR 1.48 (1.17, 1.87) |

All values are shown as hazard ratios (95% confidence interval). HR = hazard ratio; LVAD = Left Ventricular Assist Device.
95% confidence intervals for survival proportions were calculated using the Pearson-Klopper method.

**Table 2** presents the survival estimates for the study groups at each time point, which varied significantly for each (log-rank p < 0.001 for all). Survival was similar between the t-LVAD and no LVAD groups for each time point. Survival rates were lower for the d-LVAD group compared to the other two groups at each time point. **Table 2** also shows the results of the adjusted Cox proportional hazard regression model. Compared with individuals in the no LVAD group, individuals in the d-LVAD group had a significantly higher risk of mortality at each time point, with the adjusted HR ranging from 1.44 (95% CI 1.24-1.68) at 30 days to 1.18 (95% CI 1.09-1.28) at 2 years post-HT. Similarly, after risk adjustment, mortality risk was consistently higher in the d-LVAD compared with the t-LVAD groups at each time point. Causes of post-HT mortality by group in the duration of follow-up are shown in **Supplementary Table 1**. Overall, the causes did not differ significantly across groups (p = 0.45).

**Table 3** presents the results of the adjusted Cox proportional hazard regression model at each time point separately for transplants conducted before and after the UNOS allocation change. Under the old UNOS allocation policy, there was no significant difference in mortality between the t-LVAD and no-LVAD group at any time point. Conversely, compared with no-LVAD use, d-LVAD use was associated with higher mortality at each time point. Under the new UNOS allocation policy, survival decreased further in the d-LVAD group compared to the other groups, with the overall cohort (**Supplementary Figure 1**). Unlike under the old UNOS policy, risk-adjusted mortality during the new policy period was significantly higher for the d-LVAD group compared to the t-LVAD group at each time point.

**Table 3.** Analysis of Mortality by Transplant Era (2011-2018, 2018-2024)

|  | **Old UNOS Era (2011-10/18/2018; n=14,795)** |  |  | **Current UNOS Era (10/19/2018-2024; 9,826)** |  |  |
| --- | --- | --- | --- | --- | --- | --- |
| **Outcome** | **Durable LVAD vs. no LVAD** | **Temporary LVAD vs. no LVAD** | **Durable LVAD vs. Temporary LVAD** | **Durable LVAD vs. no LVAD** | **Temporary LVAD vs. no LVAD** | **Durable LVAD vs. Temporary LVAD** |
| **30-Day** | HR 1.26 (1.04, 1.52) | HR 1.18 (0.44, 3.18) | HR 1.07 (0.40, 2.89) | HR 1.69 (1.32, 2.18) | HR 0.41 (0.20, 0.82) | HR 4.14 (2.05, 8.37) |
| **90-Day** | HR 1.15 (1.00, 1.34) | HR 0.97 (0.43, 2.17) | HR 1.20 (0.53, 2.69) | HR 1.48 (1.22, 1.80) | HR 0.82 (0.55, 1.22) | HR 1.81 (1.20, 2.71) |
| **1-Year** | HR 1.22 (1.00, 1.26) | HR 1.15 (0.63, 2.10) | HR 0.98 (0.54, 1.78) | HR 1.37 (1.18, 1.59) | HR 0.79 (0.59, 1.07) | HR 1.72 (1.26, 2.36) |
| **2-Year** | HR 1.13 (1.02, 1.25) | HR 1.03 (0.59, 1.78) | HR 1.10 (0.63, 1.91) | HR 1.26 (1.05, 1.44) | HR 0.86 (0.66, 1.12) | HR 1.47 (1.12, 1.93) |

All values are shown as hazard ratios (95% confidence interval). HR = hazard ratio; LVAD = Left Ventricular Assist Device; UNOS = United Network for Organ Sharing

### Subgroup Analysis of Patients with Available Device Implant Date

A total of 4,471 patients had device implantation data available: 3,757 with durable and 714 with temporary LVAD. The characteristics of individuals with an available device implant date who use d-LVAD or t-LVAD are shown in **Supplementary Table 2**. After adjusting, the use of a d-LVAD was associated with a higher risk of 1-year post-transplant mortality compared with a t-LVAD (HR 1.32, 95% CI 0.98-1.78, p=0.072). When modeled continuously as a restricted cubic spline, **Figure 3** shows the relationship between increasing time on d-LVAD (any device) and hazard of 1-year mortality, with the median support time (493 days) serving as the reference. When compared with d-LVAD less than 2 years, being on any device for ≥2 years was associated with a 39% increased hazard of 1-year mortality (HR 1.39, 95% CI 1.15-1.68).

**Figure 3:**
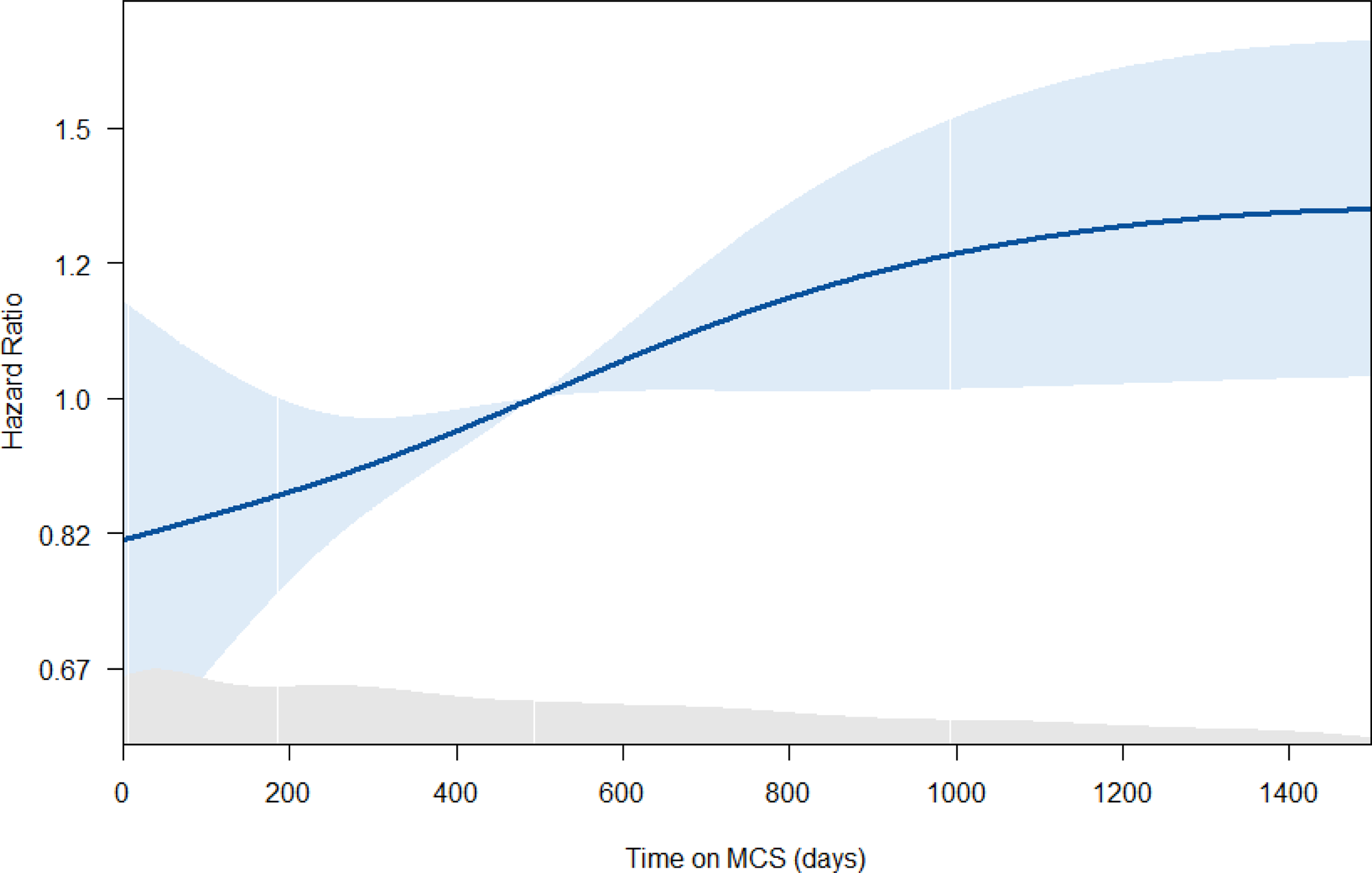
Relationship Between Time on MCS and Adjusted 1-Year Mortality Hazard Ratio. This analysis was limited to the t-LVAD and d-LVAD groups. The thick blue line represents the spline relationship between time on MCS (in days) and the hazard ratio for 1-year mortality, with the reference point being the median time on MCS value of 493 days. The thin blue shade represents the 95% CI. The grey shading at the bottom represents the proportion of the population at each time point. MCS = Mechanical Circulatory Support.

Time on LVAD (regardless of the device) was then divided into quartiles, with the Kaplan-Meier plot for each shown in **Figure 4**. Compared with the shortest duration quartile (0-184 days), increasing time on LVAD to 185-493 days (HR 1.36, 95% CI 1.01-1.84), 494-992 days (HR 1.50, 95% CI 1.11-2.01), and 993+ days (HR 1.73, 95% CI 1.30-2.31) was associated with increasing hazard for 1-year mortality.

**Figure 4:**
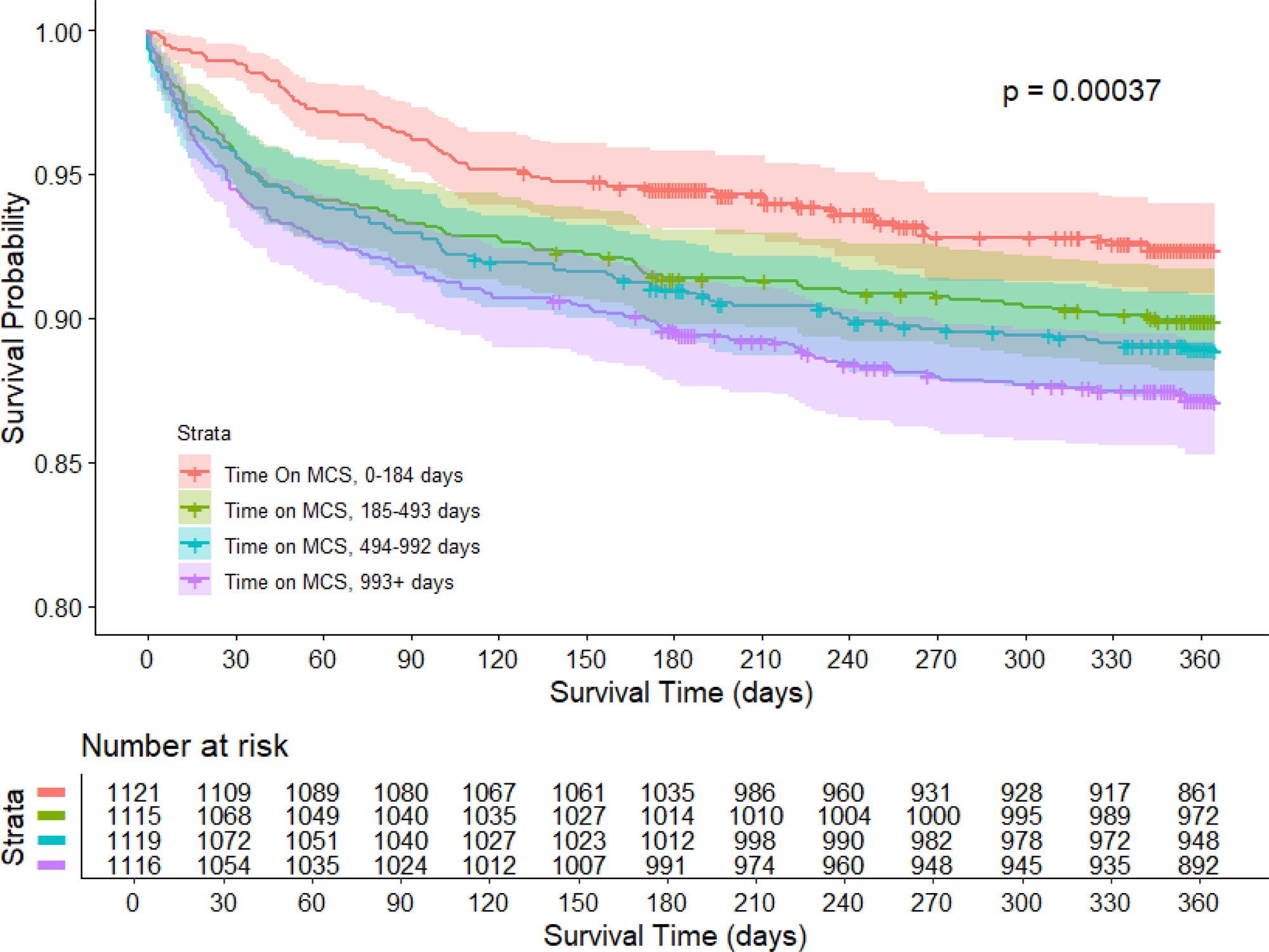
Kaplan-Meier Analysis for 1-Year Survival by Time on Mechanical Circulatory Support Category. CAT = category; MCS = mechanical circulatory support

### Subgroup Analysis of Patients with a HeartMate 3 Durable LVAD

Of the 10,729 individuals in the durable LVAD arm, 2,222 (20.7%) had received HM3 devices. The total of this subgroup cohort was 16,288 (13,170 [80.9%] with no LVAD, 2,222 [13.6%] with a HM3 LVAD, and 896 [5.5%] with a tLVAD). Compared with individuals in the no LVAD group, the hazard of 1-year mortality was higher in the HM3 group (HR 1.27, 95% CI 1.09-1.48) but not the t-LVAD group (HR 0.77, 95% CI 0.59-1.00), and higher in the HM3 group compared to the t-LVAD group (HR 1.65, 95% CI 1.23-2.21).

### Subgroup Analysis by Listing Priority

From the overall cohort of 24.795 individuals, 15,117 (61%) were high-priority listings as compared with 9,678 (39%) who were low-priority listings. Amongst the high-priority listings, death within the first year after HT occurred in 1,330 (8.8%) individuals. Compared to individuals in the no LVAD group, the hazard of 1-year mortality was higher in the d-LVAD group (HR 1.24, 95% CI 1.10-1.40), lower in the t-LVAD group (HR 0.76, 95% CI 0.58-0.99), and higher in the d-LVAD vs. t-LVAD groups (HR 1.65, 95% CI 1.25-2.16). Amongst the low-priority listings, only 5 (0.1%) individuals were on a t-LVAD and overall death within the first year after HT occurred in 814 (8.4%) individuals. Compared to individuals in the no LVAD group, the hazard of 1-year mortality was higher in the d-LVAD group (HR 1.16, 95% CI 1.00-1.35). Comparisons with the t-LVAD group were not conducted due to so few patients being in that group.

### Subgroup Analysis of Only Patients Who Received a DCD Transplant

A total of 888 (3.6%) DCD transplants were identified. Compared with individuals in the no LVAD group, the hazard of 1-year mortality was similar in both the d-LVAD group (HR 1.34, 95% CI 0.78-2.28) and the t-LVAD group (HR 1.38, 95% CI 0.51-3.73), with no significant difference observed between the d-LVAD and t-LVAD groups (HR 0.97, 95% CI 0.35-2.65). Additionally, 677 (2.9%) transplants utilizing machine perfusion were identified. There was no evidence of a difference in 1-year mortality hazard between the d-LVAD group (HR 1.49, 95% CI 0.83-2.68) or the t-LVAD group (HR 1.12, 95% CI 2.57-4.89) and the no-LVAD groups, nor between the d-LVAD and t-LVAD groups (HR 1.33, 95% CI 0.30-5.85).

## DISCUSSION

In this large, national analysis of over 24,000 adult HT recipients between 2011 and 2023, we found that post-transplant survival varied significantly by type of mechanical circulatory support at the time of transplant and the era of UNOS allocation policy. Notably, recipients bridged with durable LVADs had higher risk-adjusted mortality at every post-transplant timepoint up to 2 years compared with those bridged with temporary or no LVAD support. These findings were consistent both before and after October 2018 UNOS heart allocation policy change. Still, the mortality gap between d-LVAD and t-LVAD recipients was even more pronounced in the modern era. In contrast, survival among t-LVAD recipients was comparable to, and in some instances better than those with no LVAD support. However, survival in recipients bridged with either d-LVAD or t-LVAD was similar in transplants involving DCD and DBD with machine perfusion.

Our results suggest that the use of d-LVADs, while historically central to bridge-to-transplant therapy, may now be associated with inferior long-term outcomes, particularly under the current status-based donor organ allocation system (**Central Illustration**). The increased risk observed among d-LVAD recipients persisted even after adjustment for known confounders, including comorbidities, device type, and donor-recipient characteristics. This suggests that bridging with d-LVAD may be an independent risk factor for post-HT mortality.

A recent comparative analysis of data from the MOMENTUM 3 trial examined trial participants who were younger than 50 at the time of receiving a HeartMate 3 LVAD implant, and compared them to non-LVAD patients (<50 years old at the time of transplant listing) and showed similar two-year survival even after propensity matching (90.8% and 90.5%).[16] Importantly, at two years, freedom from death and delisting due to worsening clinical status was 90% in the LVAD group, compared to 78% in the waitlisted group after propensity score matching. One-year post-treatment, LVAD recipients experienced fewer infections requiring hospitalization (24.9% vs. 32.2%) and a lower incidence of renal dysfunction requiring dialysis (4.7% vs. 10.1%) compared to matched HT recipients in the UNOS registry. However, the LVAD cohort did experience higher rates of debilitating stroke (2.7% vs. 0.3%). The authors proposed that these findings lent support for a “LVAD first” treatment strategy with later bridging to transplant. It is noteworthy that patients enrolled in the MOMENTUM trial were a highly selected population and represent only a small fraction of the real-world patients who receive d-LVADs. In contrast, among the broader and more heterogeneous LVAD recipients captured in the UNOS registry, our findings demonstrate that post-transplant outcomes in patients bridged with d-LVADs were inferior, regardless of device type, donor retrieval method, and patient age, challenging the proposed “net prolongation of life” argument and suggesting a need for a nuanced approach to bridging decisions.

Several factors may explain the differential outcomes between recipients of d-LVAD and t-LVAD. Patients supported with d-LVADs typically undergo longer durations of support, exposing them to cumulative risks such as driveline infections, thromboembolic events, right heart failure, gastrointestinal bleeding, and allosensitization. [22] Moreover, these patients more frequently had prior cardiac surgeries, resulting in technically challenging and prolonged redo operations and bleeding risks, which may have compounded operative risks despite statistical adjustment.

The central distinction between the prior and current allocation systems lies in how stable VAD patients are prioritized. Under the previous system, status 1B patients (i.e., stable VAD recipients) could still undergo transplantation even without complications. In contrast, with the implementation of the broader 500-mile sharing policy in October 2018, the current allocation system has further limited access to transplantation for stable d-LVAD recipients, rendering transplantation uncommon in this population. Consequently, under the current system, stable d-LVAD recipients face limited transplant opportunities, with candidacy for transplantation largely contingent upon the development of a complication that increases their listing status. Indeed, most HT recipients with d-LVADs (62%) received organs while listed as UNOS status 2 or 3, indicating underlying complications that necessitated an upgrade in priority. Compounding this, right heart failure, while common in this population, does not in itself confer higher status unless it necessitates an RVAD, leading to the accumulation of morbidity and the clinical consequences of right heart failure until transplantation is ultimately performed, often as a reoperation. Accordingly, it is not unexpected that our analysis demonstrates further worsening of survival for BTT patients supported with d-LVADs in the modern allocation era.

Our subgroup analysis further supports the negative impact of prolonged LVAD exposure on post-transplant outcomes. Time on support, regardless of device type, was associated with increasing 1-year mortality in a stepwise fashion. Patients supported for more than two years had a nearly 40% higher adjusted hazard of death within the first year post-transplant. These findings are consistent with prior studies demonstrating that prolonged LVAD support is associated with adverse immunologic and end-organ effects, including sensitization, hepatic congestion, and renal dysfunction, all of which can compromise transplant candidacy and outcomes. On the other hand, patients on t-LVADs were younger and more acutely ill but may have benefited from expedited transplant under the new UNOS allocation criteria.[23] We must highlight our observation of the favorable survival of tLVAD patients, even surpassing no-LVAD recipients. Whether such a finding is a result of the reversal of cardiogenic shock with t-LVAD, tLVAD being leveraged strategically for allocation advantage, or due to unmeasured bias remains unclear, beyond the ambit of current analysis and warrants further investigation. However, we do caution against a simplistic conclusion that every patient should receive a t-LVAD before transplant.

A recent UNOS registry report evaluating DCD transplantation showed higher mortality among patients bridged with LVADs.[24] The authors also analyzed device-specific outcomes and reported similar results for patients supported with HM3 LVADs. In our analysis, we also observed higher mortality with DCD transplants among LVAD patients, regardless of the LVAD device used. The differences in findings between the two studies may be attributed to our longer study duration, larger patient cohort, more comprehensive handling of missing data, and adjustment for different covariates. Nevertheless, both studies reported nearly identical 1-year survival rates: 93.6% (no LVAD) and 88.4% (LVAD) in the UNOS study, versus 93.9% (no LVAD) and 89.5% (LVAD) in our analysis. Given the relatively small number of events, such methodological and temporal differences may contribute to variations in the 1-year hazard ratios, even though the absolute differences are modest. In summary, despite differences in timing and methodology, the results across studies trended in a similar direction.

Our findings address a central and unresolved question in the heart transplant community: In the current U.S. allocation system, should clinically deteriorating patients be bridged with a d-LVAD and wait—potentially accumulating device-related complications—before undergoing transplant, or should they proceed directly to transplant with a temporary LVAD? Durable LVADs are not without risk, and complications tend to accumulate over time. While not definitively proven, prolonged d-LVAD support may impart an adverse effect on post-transplant survival. This raises important considerations: Is there an early mortality cost associated with extended d-LVAD support? Does conditional 1-year survival differ based on the type and duration of pre-transplant LVAD? Furthermore, is long-term (e.g., 10-year) survival and complication burden more favorable with direct transplantation compared to a pathway involving prolonged d-LVAD support followed by delayed transplant? Our findings provide new insights into these critical questions and suggest that the choice and timing of LVAD strategy may have durable implications for transplant outcomes. Given the ongoing shortage of donor organs, realistically, we cannot escape d-LVAD as a bridge strategy, and our observations also lend support to opportunities in refining the allocation policy to ensure earlier and fairer access for stable d-LVAD patients as well as defining a subgroup of d-LVAD patients who might still benefit from bridging with d-LVAD for transplant in prospective studies.

### Limitations

Our study has several limitations. Despite using a comprehensive national dataset, certain granular clinical data—such as specific immunosuppressive regimens, center-level practices, or intraoperative variables—were not available. Although we adjusted for multiple covariates and used multiple imputations to handle missing data, residual confounding remains a possibility. Selection bias may exist in terms of which patients receive temporary versus durable devices, particularly considering center preferences and organ availability. Additionally, t-LVADs do carry risks of their own, and this study could not account for patients who might have died or were delisted due to t-LVAD complications, some of whom may have subsequently undergone d-LVAD implantation as a bailout strategy. Outcomes beyond 2 years were not evaluated due to the limited duration of follow-up in the t-LVAD arm. Subgroup analyses, particularly those looking at the DCD groups, were relatively small compared with the number of events. As a result, analyses are likely overfit given the factors adjusted for; therefore, results must be interpreted with caution. Lastly, unmeasured center-level factors, such as experience with temporary and durable LVAD devices and local listing practices, may influence outcomes but were not captured in our analysis.

## CONCLUSIONS

Durable LVAD support at the time of heart transplantation is associated with significantly higher post-transplant mortality than temporary LVAD or no LVAD use. This disparity has widened in the current UNOS allocation era. Time on (durable) LVAD support and transplant at higher priority status are independent risk factors for worse post-transplant survival in patients with d-LVADs, reinforcing the importance of minimizing LVAD duration when feasible. Our findings suggest that clinical pathways and organ allocation policies should be reevaluated to minimize prolonged durable LVAD support where feasible and prioritize earlier transplant when clinically appropriate.

## Data Availability

All data is publicly available upon request by UNOS

## Acknowledgement

We would like to thank Mary Ann Citraro, Suzanne Turner, and Charlene Surace, Department of Cardiology Graphics & Design at Cleveland Clinic, for assistance with the central illustration.

## Perspectives

### What is new?

The routine use of durable LVAD as a bridge to transplant, especially beyond two years, worsens long-term post-transplant survival. Early transplantation with temporary LVAD may offer superior outcomes and should be considered where feasible.

### What are the clinical implications?

Future donor allocation policy and research should address the observed survival disadvantage associated with prolonged durable LVAD use. The development of dynamic risk stratification tools that incorporate support duration, device type, and device-related complications is essential for guiding the optimal timing of transplantation and improving long-term outcomes.

---

**Supplementary Table 1.** Causes of mortality in study groups 2-year post-transplant.

| Cause of Death | No LVAD<br>(n=1,393/13,170,<br>10.6%) | Durable LVAD<br>(n=1,406/10,729,<br>13.1%) | Temporary LVAD<br>(n=78/896, 8.7%) |
| --- | --- | --- | --- |
| Cardiovascular | 231 (16.6%) | 199 (14.2%) | 17 (21.8%) |
| Cerebrovascular | 91 (6.5%) | 119 (8.5%) | 5 (6.4%) |
| Graft Failure | 193 (13.9%) | 206 (14.7%) | 12 (15.4%) |
| Hemorrhagic | 39 (2.8%) | 53 (3.8%) | 2 (2.6%) |
| Infection | 266 (19.1%) | 256 (18.2%) | 13 (16.7%) |
| Other/Not specified | 476 (34.2%) | 475 (33.8%) | 25 (32.1%) |
| Pulmonary | 91 (6.5%) | 96 (6.8%) | 4 (5.1%) |
| Unknown | 6 (0.4%) | 2 (0.1%) | 0 (0%) |

**Supplementary Table 2.** Characteristics of the study cohort with an available device implantation date.

| Characteristic | Total Cohort<br>(n=4,471) | Temporary LVAD<br>(n=714) | Durable LVAD<br>(n=3,757) | p-value |
| --- | --- | --- | --- | --- |
| **Recipient Characteristics** |  |  |  |  |
| Age, years | 56 (46, 63) | 56 (46, 64) | 56 (46, 63) | 0.8 |
| Men | 3,502 (78%) | 578 (81%) | 2,924 (78%) | 0.063 |
| Race |  |  |  | <0.001 |
| Asian | 127 (2.8%) | 27 (3.8%) | 100 (2.7%) |  |
| Black | 1,212 (27%) | 167 (23%) | 1,048 (28%) |  |
| Other | 492 (11%) | 107 (15%) | 385 (10%) |  |
| White | 2,640 (59%) | 413 (58%) | 2,227 (59%) |  |
| BMI, kg/m<sup>2</sup> | 29.3 (25.8, 33.0) | 26.7 (23.5, 30.6) | 29.8 (26.3, 33.3) | <0.001 |
| HF Etiology |  |  |  | 0.12 |
| CHD | 36 (0.8%) | 4 (0.6%) | 32 (0.9%) |  |
| DCM | 2,771 (62%) | 433 (61%) | 2,338 (62%) |  |
| HCM | 23 (0.5%) | 6 (0.8%) | 17 (0.5%) |  |
| ICM | 1,371 (31%) | 200 (28%) | 1,171 (31%) |  |
| Other | 215 (4.8%) | 58 (8.1%) | 157 (4.2%) |  |
| RCM | 55 (1.2%) | 13 (1.8%) | 42 (1.1%) |  |
| History of diabetes mellitus | 1,409 (32%) | 230 (32%) | 1,179 (31%) | 0.7 |
| Prior cardiac surgery | 2,491 (57%) | 174 (25%) | 2,317 (63%) | <0.001 |
| Ventilator at transplant | 17 (0.4%) | 11 (1.5%) | 6 (0.2%) | <0.001 |
| Creatinine, mg/dL | 1.16 (0.94, 1.41) | 1.10 (0.89, 1.42) | 1.17 (0.96, 1.41) | <0.001 |
| Bilirubin, mg/dL | 0.6 (0.4, 0.9) | 0.8 (0.5, 1.4) | 0.6 (0.4, 0.8) | <0.001 |
| PRA, % | 0 (0, 26) | 0 (0, 23) | 0 (0, 26) | 0.3 |
| PCWP, mmHg | 13 (9, 20) | 22 (15, 28) | 12 (8, 18) | <0.001 |
| CO, L/min | 4.70 (3.90, 5.59) | 4.31 (3.40, 5.60) | 4.77 (4.00, 5.58) | <0.001 |
| Old Listing Status |  |  |  | <0.001 |
| 1A | 108 (2.4%) | 2 (0.3%) | 106 (2.8%) |  |
| 1B | 18 (0.4%) | 1 (0.1%) | 17 (0.5%) |  |
| New Listing Status |  |  |  | <0.001 |
| 1 | 238 (5.5%) | 67 (9.4%) | 171 (4.7%) |  |
| 2 | 1,431 (33%) | 592 (83%) | 839 (23%) |  |
| 3 | 1,468 (34%) | 25 (3.5%) | 1,443 (40%) |  |
| 4 | 1,207 (28%) | 27 (3.8%) | 1,180 (32%) |  |
| 6 | 1 (<0.1%) | 0 (0%) | 1 (<0.1%) |  |
| **Donor Characteristics** |  |  |  |  |
| Donor age, years | 33 (25, 41) | 32 (25, 40) | 33 (25, 41) | 0.3 |
| Donor Male | 3,367 (75%) | 553 (77%) | 2,814 (75%) | 0.15 |
| Donor BMI, kg/m<sup>2</sup> | 27 (24, 32) | 27 (24, 31) | 27 (24, 32) | 0.057 |
| Donor history of diabetes mellitus | 208 (4.7%) | 27 (3.8%) | 181 (4.9%) | 0.2 |
| Donor history of hypertension | 721 (16%) | 110 (16%) | 611 (16%) | 00.6 |
| **Transplant Characteristics** |  |  |  |  |
| Ischemic time, h | 3.52 (2.87, 4.18) | 3.63 (3.00, 4.24) | 3.50 (2.85, 4.17) | 0.002 |
| Donor-recipient PHM ratio | 1.09 (1.00, 1.21) | 1.12 (1.02, 1.22) | 1.09 (0.99, 1.20) | <0.001 |
| Recipient-Donor Mismatch | 795 (18%) | 135 (19%) | 660 (18%) | 0.4 |
| Follow-up Time, y | 2.58 (1.02, 4.02) | 1.05 (0.91, 3.06) | 2.93 (1.13, 4.05) | <0.005 |
| Geographic Region |  |  |  | <0.001 |
| Midwest | 1,154 (26%) | 120 (17%) | 1,034 (28%) |  |
| Northeast | 1,084 (24%) | 163 (23%) | 921 (25%) |  |
| South | 1,513 (34%) | 283 (40%) | 1,230 (33%) |  |
| West | 720 (16%) | 148 (21%) | 572 (15%) |  |

Data are presented either as n (%) or median (25<sup>th</sup>, 75<sup>th</sup> percentile) and compared using either Pearson's chi-squared test or Wilcoxon rank sum test, respectively. BMI = body mass index; CHD = congenital heart disease; CO = cardiac output; DCM = dilated cardiomyopathy; HCM = hypertrophic cardiomyopathy; ICM = ischemic cardiomyopathy; PCWP = pulmonary capillary wedge pressure; PHM = predicted heart mass; PRA = panel-reactive antibody; RCM = restrictive cardiomyopathy

**Supplementary Figure 1.**
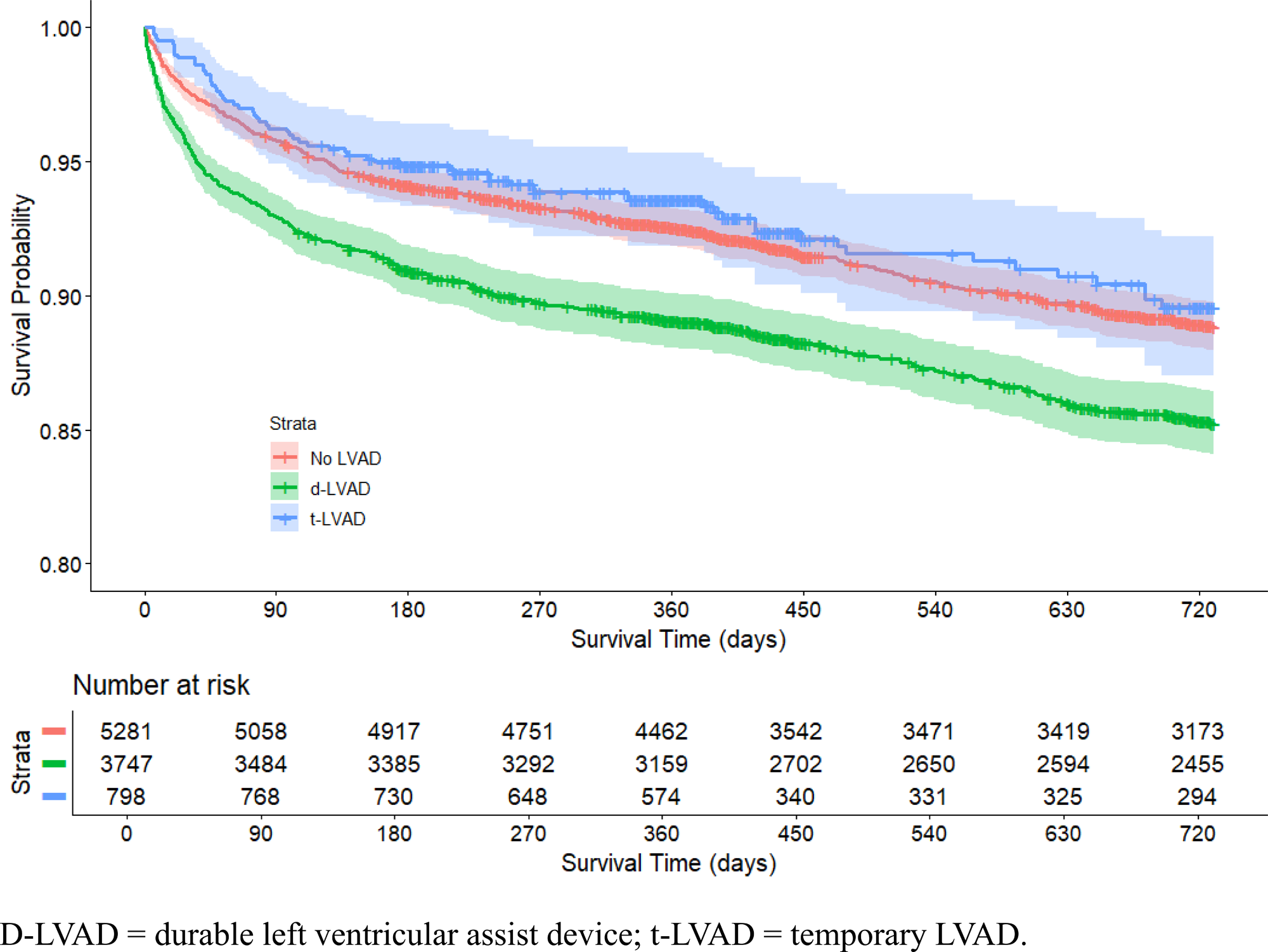
Kaplan-Meier Analysis for 2-Year Survival Under the New UNOS Allocation System.

**Central Illustration:**
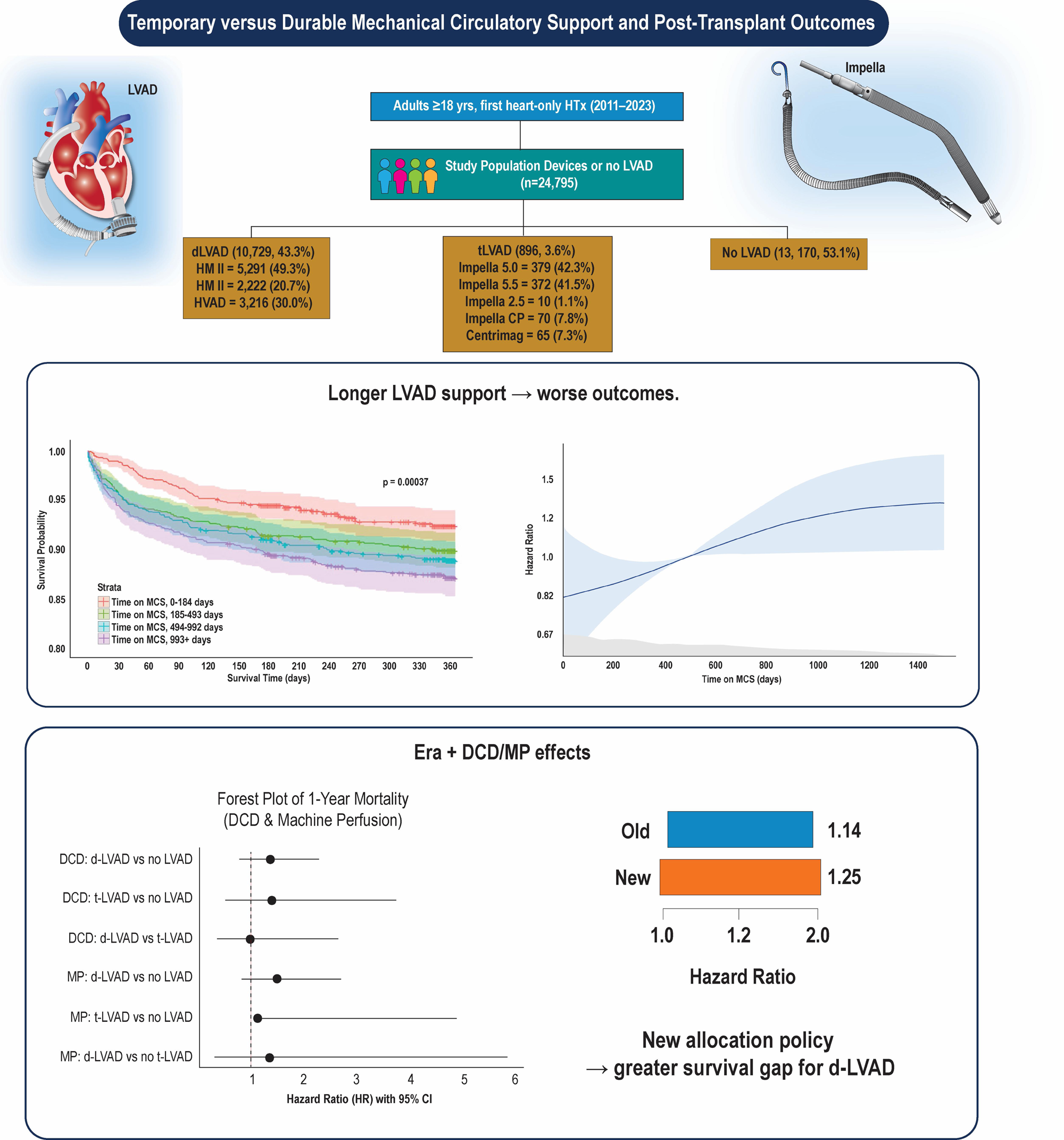
Temporary versus durable mechanical circulatory support and post-transplant outcomes. DCD: donation after circulatory death; HTx: heart transplant; LVAD: left ventricular assist device; d-LVAD: durable LVAD; t-LVAD: temporary LVAD; MCS: mechanical circulatory support; MP: machine perfusion.

